# Fractal dimension distributions of resting-state EEG improve detection of dementia and Alzheimer’s disease compared to traditional fractal analysis

**DOI:** 10.1101/2024.06.10.24308727

**Authors:** Keith J. Yoder, Geoffrey Brookshire, Ryan Glatt, David A. Merrill, Spencer Gerrol, Colin Quirk, Ché Lucero

## Abstract

Across many resting-state electroencephalography (EEG) studies, dementia is associated with changes to the power spectrum and fractal dimension. Here we describe a novel method to examine changes in fractal dimension over time and within frequency bands. This method, which we call Fractal Dimension Distributions (FDD), combines spectral and complexity information. In this study, we illustrate this new method by applying it to resting-state EEG data recorded from patients with subjective cognitive impairment (SCI) or dementia. We compared the performance of FDD with the performance of standard fractal dimension metrics (Higuchi and Katz FD). FDD revealed larger group differences detectable at greater numbers of EEG recording sites. Moreover, linear models using FDD features had lower AIC and higher R^2^ than models using standard full time-course measures of fractal dimension. FDD metrics also outperformed the full time-course metrics when comparing SCI with a subset of dementia patients diagnosed with Alzheimer’s disease (AD). FDD offers unique information beyond traditional full time-course fractal analyses and may help to identify AD dementia and non-AD dementia.

## 1. Introduction

Alzheimer’s Disease (AD) is the leading cause of dementia and the sixth most common cause of death in the United States [1]. The risk of developing AD grows with age; about 10.7% of adults aged 65 or older have Alzheimer’s dementia, and that percentage climbs to 33.2% for those aged 85 or older [2]. AD is ultimately fatal, with no cure and only symptomatic treatments [3]. The annual incidence of AD is projected to double by 2050 due to increasing life expectancies [4], necessitating new methods to treat and test for the disease.

Most AD patients first experience a stage of mild cognitive impairment (MCI), where memory loss and other cognitive changes are pronounced enough to register on clinical assessments, but not so extreme as to interfere with independent daily functioning. Before being diagnosed with MCI, some patients report worsening cognitive abilities despite scoring within healthy ranges on clinical assessments[5]; this condition is referred to as subjective cognitive impairment (SCI). AD is primarily diagnosed through clinical cognitive assessments [6], which may not always catch early-stage cognitive impairment or distinguish between different forms of dementia [7]. Historically, AD diagnoses were only confirmed post-mortem through an autopsy. There has been a push to incorporate more objective biomarker-based tests into clinical practice and research [8], but these tests are often invasive (e.g. a lumbar puncture) or expensive (e.g. positron emission tomography). While new plasma biomarkers have demonstrated excellent accuracy at detecting amyloid and tau pathology [9], these methods can take time to compute and may not be strongly associated with cognitive impairment. A growing body of research positions the electroencephalogram (EEG) as a fast, inexpensive, non-invasive biomarker for AD [10].

AD patients present with a number of EEG abnormalities, one of which is reduced signal complexity [11–13]. Complexity in brain signals arise from interacting neural circuits operating over multiple spatial and temporal scales. A variety of complexity methods have been previously used, including entropy, Hurst exponent, correlation dimension, and fractal dimension (reviewed in [12]). Fractal dimension (FD) is a nonlinear measure that expresses how the details of a self-similar form are altered by the scale at which they are measured[14]. In this study, we utilize both the Katz Fractal Dimension (KFD) and the Higuchi Fractal Dimension (HFD) methods. KFD calculates the fractal dimension by comparing distances along the waveform [15], whereas HFD approximates the box-counting dimension in time-series data by repeatedly downsampling a waveform and comparing the length of the subsampled waveforms to the downsampling factor [16]. Though KFD tends to underestimate FD, KFD is sometimes better at discriminating between different brain states than HFD [17].

Diminished HFD and KFD are observed in the EEG and magnetoencephalography (MEG) of individuals with AD and dementia [18–22]. While EEG signal complexity drops after age 60 even in the absence of disease, AD is associated with a decrease in FD beyond what is observed in healthy, age-matched controls [20]. HFD and KFD have been used in conjunction with machine learning algorithms to distinguish between AD and healthy controls with high accuracy and specificity, making them a prime candidate for an EEG-based AD biomarker [23–25]. These algorithms can be improved by computing FD within distinct frequency bands (e.g. delta, theta, alpha) rather than in broadband EEG [24,26]. For instance, Nobukawa and colleagues [21] found that the difference in a modified HFD between AD and healthy controls was greater at higher frequencies than lower frequencies.

We propose that the predictive capability of EEG could be further improved by analyzing the distribution FD over the course of an EEG session. A common implementation of HFD and KFD algorithms compute a single value summarizing the entire time series, meaning any changes in FD over time are lost. Some previous work calculated FD within moving windows but discarded information about potential changes in FD over time by averaging the resulting values together [22,25]. However, the distribution of FD over time may contain valuable information. Here we develop a new technique called Fractal Dimension Distributions (FDD). Rather than assessing FD using the full EEG time-course, FDD slides a moving window across the full time-course, computes FD (HFD or KFD) within each window, and then summarizes the distribution of FD values across time-windows (e.g. mean, standard deviation). We provide an initial demonstration of this approach by calculating FDD in participants with SCI or dementia. We then evaluate whether FDD carries more information about dementia than traditional full-time-course metrics of FD.

## 2. Materials and Methods

### 2.1. Study population

All patients were adults over the age of 55 who visited a specialty memory clinic (Pacific Brain Health Center in Santa Monica, CA) for memory complaints. Adults were evaluated by a dementia specialist during their visit. Evaluations included behavioral testing and EEG recordings. Patients with SCI or dementia were selected retrospectively by reviewing charts for patients seen between July 2018 and February 2021. Full data was available from 148 adults (91 female, Age M=71.3 years, SD=7.5). Groups were divided into adults diagnosed with SCI (N=97, 59 [60.8%] female, Age M=70.2, SD=7.1) or dementia (N=51, 32 [62.7%] female, Age M=73.7, SD=7.8). Within the dementia group, 38 individuals were diagnosed with AD (26 [68.4%] female, Age M=74.2, SD=7.1). The remaining individuals were diagnosed with Lewy body dementia (n=4), vascular dementia (n=2), frontotemporal dementia (n=2), Parkinson’s disease (n=2), or unknown (n=3). All procedures aligned with the Helsinki Declaration of 1975 and were approved by the Institutional Review Board at the St. John’s Cancer Institute. All patients provided informed consent.

### 2.2. Clinical diagnosis

Patient diagnosis was based on consensus of a panel of board-certified dementia specialists using the 2011 guidelines proposed by the National Institute of Aging and the Alzheimer’s Association (NIA-AA). Diagnoses utilized standard clinical methods for neurological examinations, cognitive testing (MMSE [27] or MoCA [28]), clinical history (e.g. depression, diabetes, head injury, hypertension), and laboratory testing (e.g. thyroid stimulating hormone levels, rapid plasma reagin testing, vitamin B-12 levels). Cognitive impairment was diagnosed on the basis of the MMSE (or MoCA scores converted to MMSE [29]). MCI was diagnosed according to the criteria established Langa & Levine [30], and distinguished from dementia on the basis of preserved functional abilities and independence together with a lack of significant impairment in occupational or social functioning. SCI was diagnosed based on subjective complaints without evidence of MCI.

### 2.3. EEG collection

EEG data were recorded using a 19-channel eVox System (Evoke Neuroscience) at the Pacific Neuroscience Institute. Electrodes were positioned in a cap according to the international 10-20 system (Fp1, Fp2, F7, F3, Fz, F4, F8, T7, C3, Cz, C4, T8, P7, P3, Pz, P4, P8, O1, and O2). Data were collected at 250 Hz while patients completed two resting-state recordings — five minutes each of eyes closed and eyes open — and a 15 minute Go/No-Go task. For this study, we used only the eyes-closed resting-state data.

### 2.4. EEG preprocessing

EEG data were re-referenced offline to the average of all channels. Jump artifacts were identified as global field power more than 10 standard deviations from the mean, over a maximum of 10 sequential samples. These artifacts were removed and replaced with a linear interpolation between the nearest samples that were not contaminated by jump artifacts, computed separately for each channel. Ocular artifacts were removed with the aid of Independent Component Analysis (ICA). 18 ICA components were extracted, then compared to a template of stereotypical ocular artifacts. After the ocular artifact was removed, ICs were projected back into sensor space.

For the broadband analysis, we filtered data with a 1-50 Hz zero-phase finite impulse response bandpass to help attenuate line noise. In the banded analysis, we applied separate bandpass filters for the delta (1-4 Hz), theta (4-8 Hz), alpha (8-13Hz), beta (13-30 Hz), and gamma (30-50 Hz) frequency bands. After filtering, we segmented the data into 1 s duration epochs, with a 0.5 s overlap.

### 2.5. Fractal dimension

We calculated KFD and HFD using the AntroPy package (version 0.1.4). Katz’s method[15] calculates the sum - i.e. length (*L*) - and average (*a*) Euclidean distance between successive points in the time series, as well as the maximum distance from the first point in the time series to any other point in the time series (*d*). The fractal dimension of the time series (*FD*) is then calculated as:

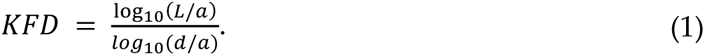

Thus, Katz’s method relies only on the raw time series itself. However, Higuchi’s method [16] begins by subsampling a time-series across progressive smaller time scales, then examining how the length of time-series is related to the scaling. Given a time series *X* ∶ {1, …, *N*} → ℝ with *N* sample points and *K_max_* ≥ 2, Higuchi’s method begins by first calculating length of a curve, 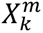, across a range of values based on initial time (*m*) and interval time (*k*). This length, *L_m_*(*k*) is defined as:

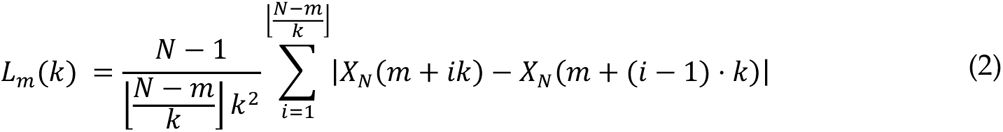

for each *m* ∈ {1, …, *k*} and *k* ∈ {1, …, *k_max_*}. Then, the length *L*(*k*) is the average:

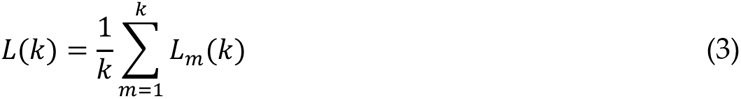

and HFD is the slope of the best-fit line through points 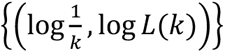.

Thus, HFD depends on both the number of sample points (N) and the parameter *k*_max_, which sets the upper limit on the number of time intervals. Some early work suggested using *k*_max_= 6 for time series with 40-1000 points [31]. Other approaches suggest calculating HFD across multiple *k*_max_ values and identifying the value of *k*_max_ at which HFD plateaus [32,33]. However, HFD is not guaranteed to plateau. Recently, Wanliss & Wanliss demonstrated that an optimal *k*_max_ can be estimated algorithmically based on the length of the time series [34]. The proposed algorithm uses two sinusoids and three pairs of parameters with empirically derived distributions. We used this algorithm to estimate the optimal *k*_max_ for the length of the full time-course, and for 1 s windows (Supplementary Materials). This indicated *k*_max_ = 108 for the full sample and *k*_max_ = 25 for 1 s windows.

#### 2.5.1. Full time-course fractal dimension

We calculated the full time-course fractal dimension by applying Katz’s method or Higuchi’s method (with *k*_max_ = 108) to the entire resting-state EEG recording. This analysis reflects the way prior studies computed FD [18,24].

#### 2.5.2. Fractal dimension distributions (FDD)

For the fractal dimension distributions, we first segmented the data using moving windows 1 s moving windows, with a 0.5 s overlap (see 3.6 for discussion of other window sizes). Within each window, we then extracted the KFD or HFD (*k*_max_ = 25) within each window. Finally, we summarized the distribution of KFD or HFD values across windows using the mean and standard deviation.

### 2.6. Group differences

We sought to compare fractal dimension measures between the SCI and dementia. We used threshold-free cluster enhancement (TFCE) to estimate the difference at each channel [35,36]. TFCE is a non-parametric technique that computes cluster-level statistics across a range of cluster forming thresholds. The channel-level TFCE statistic incorporates both the strength of the difference at that channel and the spatial extent of any neighboring clusters that exist in the data. This approach produces one TFCE value per channel. TFCE values can then be calculated for data that has had labels permuted. The final TFCE-adjusted t-statistics is thus controlled for multiple comparisons across channels. We used 10,000 permutations as implemented in the permutation_cluster_test function in the MNE python library (version 1.0.0). Positive and negative difference were calculated separately, using a step size of 0.2 Group differences were then visualized by projecting the t-statistics to the scalp with a bilinear interpolation in MNE, and individually significant channels were identified using alpha corresponding to corrected p < .05. In order to understand whether non-AD dementia and AD might be associated with different patterns of FD, we repeated this analysis comparing SCI to AD.

### 2.7. Logistic regressions

The TFCE analyses test whether FD metrics in individual channels carry information about dementia. As a complementary analysis, we used logistic regressions to test how information can be combined across channels to predict dementia (or AD). We regressed cognitive status on FD metrics in all channels, using either full time-course FD or FDD features. All models included age as a covariate. Fractal scores across channels were correlated (see Supplementary Figures 1-3), so we used regularized Least Absolute Shrinkage and Selection Operator (LASSO) regressions. For each set of predictors, a series of logistic regressions were fit, with L1 penalty values (alpha) ranging from 0 to 10 in 0.05 step increments. The model with the lowest Akaike Information Criterion (AIC) was selected for that set of predictors.

We assessed model fit using a chi-square likelihood ratio test (LRT) comparing models with fractal features to a model with age as the only predictor. The LRT is inappropriate to directly compare models using full time-course FD to models using FDD features since they are non-nested. Instead, we used AIC, with lower AIC values indicating a better model. For each model we also calculated Tjur’s coefficient of determination to obtain pseudo-R^2^, which reflects how well the model separates the two classes of patients, with 0 reflecting no separation and 1 reflecting complete separation [37].

## 3. Results

Before comparing traditional full time-course FD with the novel FDD metric, we established a baseline model, using age as the only predictor. This model was a better fit to the data than an intercept-only null model (*X^2^*(1)=7.59, p=.006). It served as a reference when conducting likelihood ratio tests for models using fractal dimension values.

### 3.1. FDD differentiates dementia better than full time-course fractal dimension

First, we tried to replicate previous studies by looking for differences in full time-course HFD and KFD between SCI and dementia [18]. In the broadband data, group differences in full time-course HFD and full time-course KFD were in the expected direction, but did not reach statistical significance. Three electrodes did show a trend towards lower HFD in the Dementia group (Figure 1; O1 t = −1.37, p = .099; Pz t = −1.40, p = .099, and P3 t = −1.44, p = .089).

**Figure 1.**
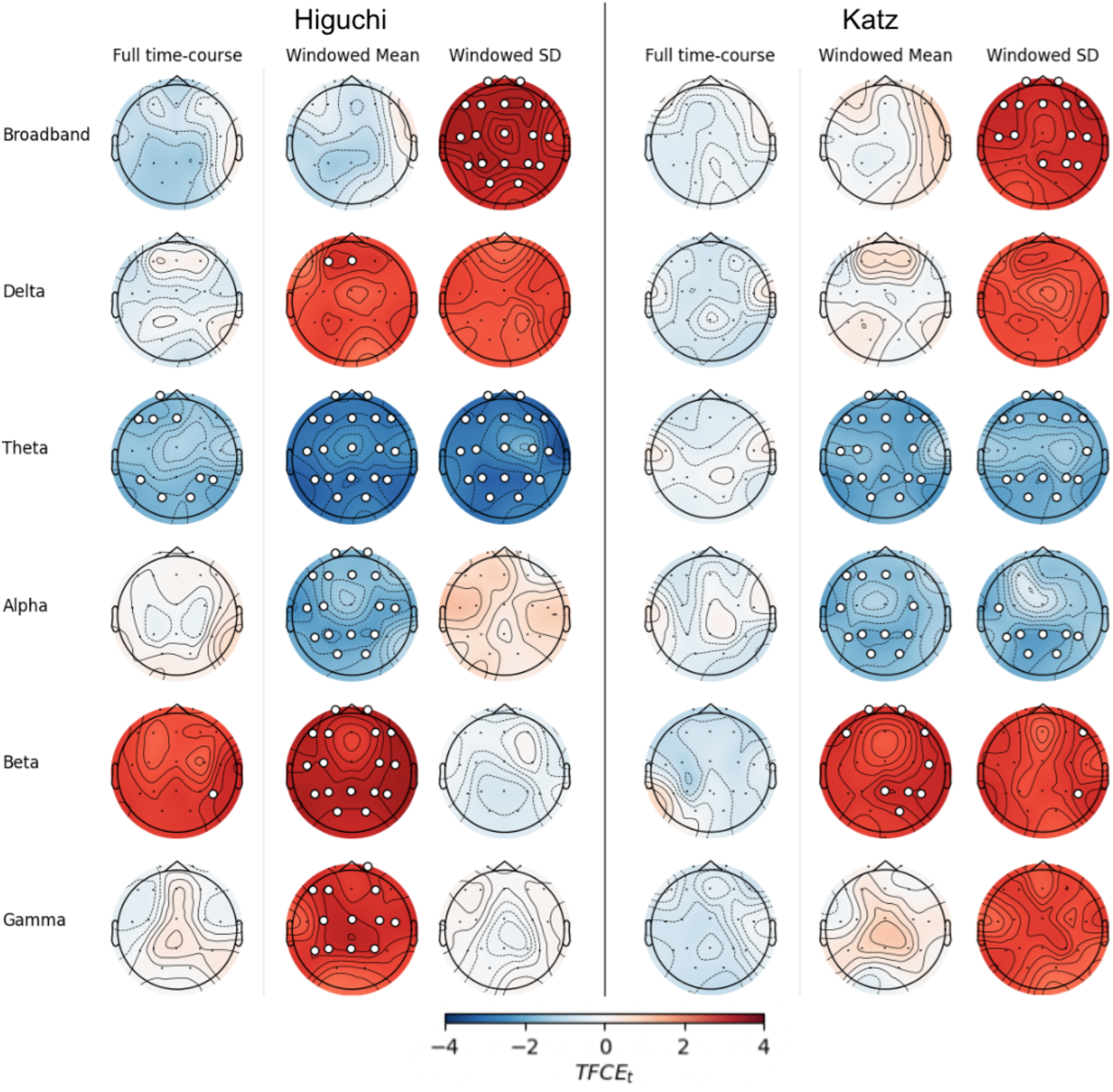
EEG fractal values differ between dementia and SCI groups. Scalp plots show differences (Dementia - SCI) calculated using threshold-free cluster enhancement (TFCE) for Higuchi fractal dimension (left) or Katz fractal dimension (right). Electrodes with significant differences (TFCE FWEp < .05) are marked with white circles.

We next examined the FDD features by calculating the mean and standard deviation of HFD and KFD across windows. In the broadband data, the mean of windowed HFD did not significantly differ between groups. In contrast, the standard deviation of windowed HFD was significantly higher in the Dementia group at every electrode (smallest difference at T8, t=1.92, p=.020, largest difference at T7, t=2.89, p<.001). Further, the standard deviation of windowed KFD was significantly higher in the Dementia group at all but five electrodes (O1, O2, P3, P7, Cz; all p > .08).

A logistic regression model using full time-course HFD showed better performance than a model using only patients’ age, and both of these were outperformed by a model using FDD features (Table 1; ΔAIC = −16.7, ΔR^2^ = .16). Similarly, a model using FDD features based on KFD outperformed a model using the full time-course KFD (Table 2; ΔAIC = −12.7, ΔR^2^ = .24). Interestingly, the lowest AIC across models using full time-course KFD was a model in which LASSO regularization set all channel coefficients to 0, retaining only age; this indicates that KFD obtained from the full time-course did not contain enough unique information relating to dementia to be included in the model.

**Table 1.** Summary of models distinguishing between dementia and SCI using Higuchi fractal information.

| Band | Fractal features | AIC | Pseudo R<sup>2</sup> | L1 penalty | n<sub>params</sub> | X<sup>2</sup> | p |
| --- | --- | --- | --- | --- | --- | --- | --- |
| Broad-band | Full time-course | 180.3 | 0.290 | 1.60 | 11 | 24.74 | **0.0033** |
|  | FDD | 163.6 | 0.446 | 1.85 | 6 | 47.40 | **0.0111** |
| Delta | Full time-course | 178.8 | 0.425 | 0.25 | 19 | 42.20 | **0.0006** |
|  | FDD | 176.1 | 0.397 | 1.90 | 19 | 44.91 | **0.0003** |
| Theta | Full time-course | 174.9 | 0.269 | 1.70 | 6 | 20.13 | **0.0005** |
|  | FDD | 159.6 | 0.592 | 0.85 | 23 | 69.44 | **0.0000** |
| Alpha | Full time-course | 182.0 | 0.262 | 1.85 | 6 | 13.04 | **0.0111** |
|  | FDD | 166.6 | 0.644 | 0.75 | 25 | 66.42 | **0.0000** |
| Beta | Full time-course | 179.5 | 0.240 | 2.15 | 7 | 17.57 | **0.0035** |
|  | FDD | 173.0 | 0.377 | 1.90 | 14 | 38.05 | **0.0002** |
| Gamma | Full time-course | 184.4 | 0.298 | 0.60 | 14 | 26.65 | **0.0087** |
|  | FDD | 184.7 | 0.220 | 4.75 | 5 | 8.33 | **0.0396** |

The number of non-zero parameters in the model (n<sub>params</sub>) including the intercept. The X<sup>2</sup> statistic and p-value reflect the likelihood ratio test compared to a model with age as the only predictor. FDD: Fractal dimension distributions.

**Table 2.**
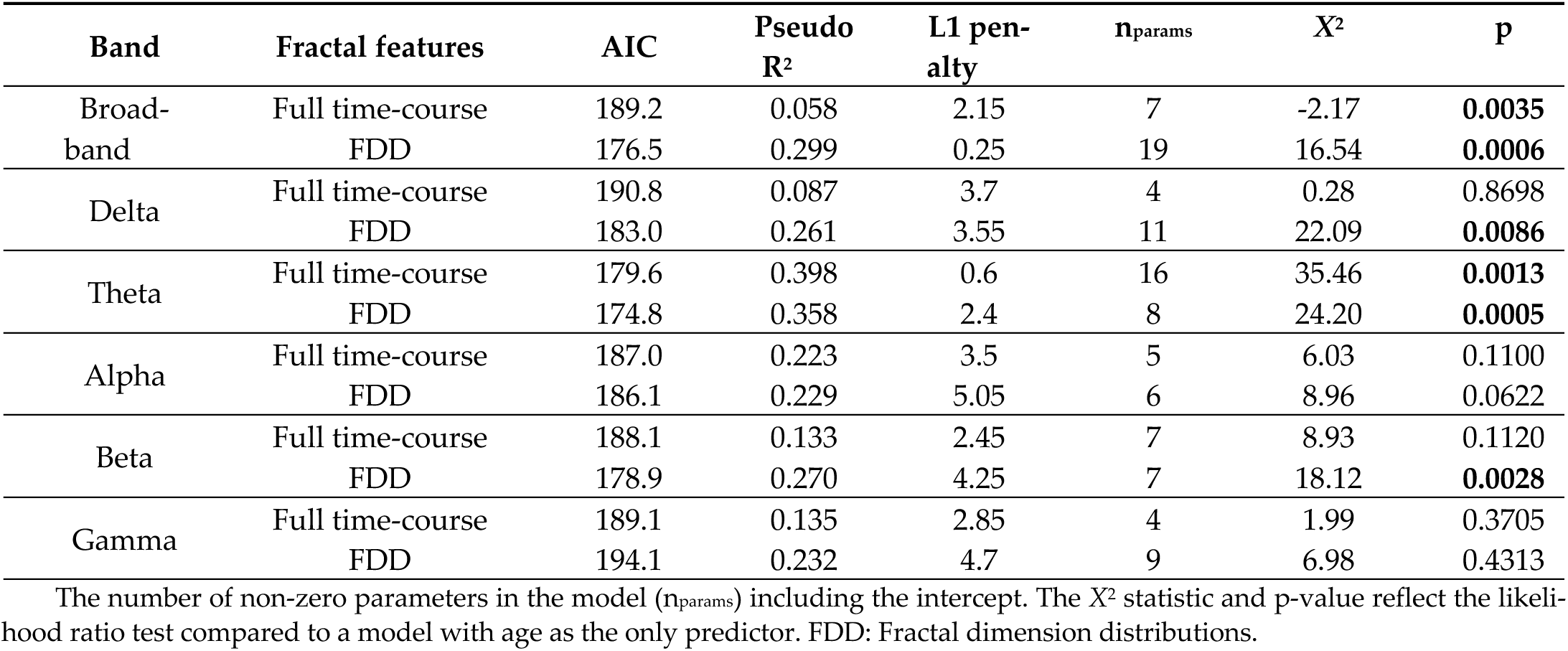
Summary of models distinguishing between dementia and SCI using Katz fractal information.

### 3.2. FDD is more informative than full time-course fractal dimension in most frequency bands

We next compared fractal dimensions between SCI and dementia within five frequency bands, calculating either the full time-course FD or FDD (Figure 1). In every band, more scalp locations showed significant group differences using Higuchi FDD calculated from full time-course HFD, particularly the windowed mean (Figure 1, left). There were no significant differences in any band using the full time-course KFD (all *p*s > 0.187). Using the Katz FDD, however, several clusters of electrodes showed significant group differences in the theta, alpha, and beta bands.

In order to visualize the extent to which FDD and full time-course FD discriminate between SCI and dementia, we plotted the absolute TFCE-t statistic averaged across channels for the full time-course FD and FDD mean and standard deviation. Figure 2 shows these values for HFD and KFD (Figure 2) for each frequency band. We then subtracted the full time-course FD values from the corresponding FDD metrics to obtain a numerical measure of the increase or decrease in the group difference at each channel - positive values indicate a larger difference using FDD features compared to full time-course FD features. We plot these relative TFCE values in each frequency band for HFD and KFD (Figure 2).

**Figure 2.**
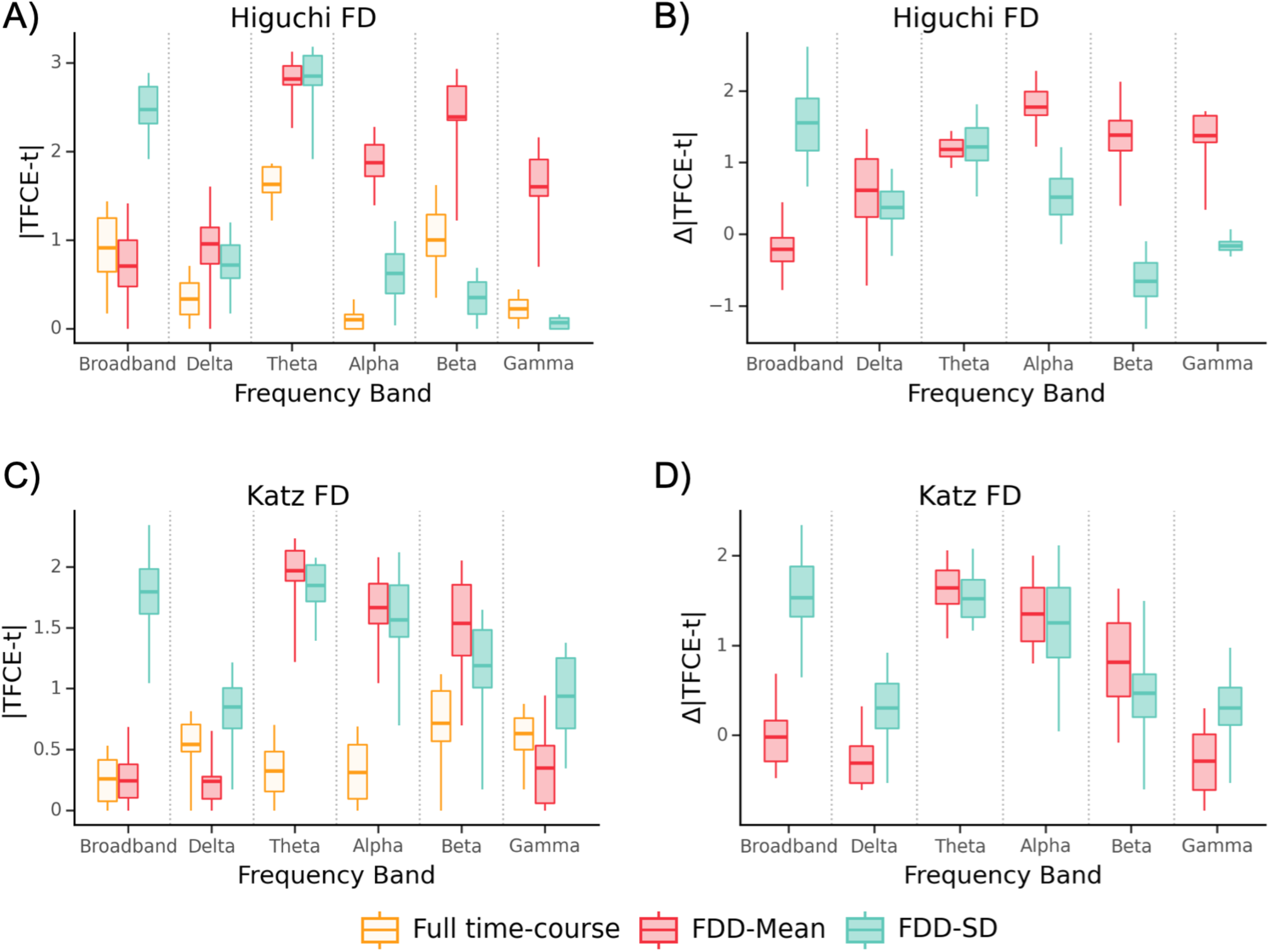
Comparison of group differences for Dem-SCI with full time-course and FDD features. Absolute TFCE-t statistic averaged across channels for group differences (Dem - SCI) in each frequency band using Higuchi (A) or Katz FD (C). Difference between FDD metrics and full time-course FD in each frequency band (B and D). Box outlines show the interquartile range, central line shows the median, and whiskers extend to minimum and maximum value.

As in the broadband analysis, we then used logistic LASSO regressions to test whether using FDD features leads to more accurate models than full time-course FD. For both Katz and Higuchi methods, models using FDD outperformed models using full timecourse FD, though the effect was stronger for HFD (Table 1; average ΔAIC = −7.9, ΔR^2^ = .15) than KFD (Table 2; average ΔAIC = −3.5, ΔR^2^ = .08). The logistic regression using FDD with Higuchi in the delta band had lower AIC but did not have higher R^2^ than the model using full time-course HFD (ΔAIC = −2.7, ΔR^2^ = -.03). In contrast, regressions using Higuchi FDD features were universally better in the theta (ΔAIC = −15.3, ΔR^2^ = .32), alpha (ΔAIC = −15.4, ΔR^2^ = .38), and beta (ΔAIC = −6.5, ΔR^2^ = .14) bands. The model with full time-course HFD had a slightly lower AIC than the model with FDD in the gamma band (ΔAIC = 0.3, ΔR^2^ = −0.08).

With Katz’s method, models using FDD had lower AIC and larger R^2^ than models using full time-course KFD in delta (ΔAIC = −7.8, ΔR^2^ = .17) and beta bands (ΔAIC = −9.2, ΔR^2^ = .14). The model using FDD features had lower AIC and slightly larger R^2^ in the alpha band (ΔAIC = −0.9, ΔR^2^ = .01). In the theta band, the model using FDD features produced a smaller AIC, but worse R^2^ (ΔAIC = −4.7, ΔR^2^ = -.04). As with HFD, in the gamma band, the model using full time-course KFD outperformed the model using FDD (ΔAIC = 5.0, ΔR^2^ = .10).

Across all models comparing Dementia and SCI, the two with the lowest AIC and highest R^2^ used Higuchi FDD features in the alpha band (AIC=166.6, R^2^ = 0.644, X^2^(25)=66.42, p < .001) or theta band (AIC=159.6, R^2^ = 0.592, X^2^(23)=69.44, p < .001).

### 3.3. FDD differentiates Alzheimer’s disease better than full time-course fractal dimension

The previous analyses demonstrate that the distribution FD carries additional information about dementia beyond the information carried by standard full time-course FD. Next, we tested whether this distributional information also helps to specifically distinguish between SCI and dementia due to Alzheimer’s disease (AD). There were no significant differences between SCI and AD in full time-course HFD or KFD when estimated from broadband data (Figure 3; all *p* > .3). There were also no significant differences in mean HFD or mean KFD at any electrode (all *p* > .2). However, KFD SD was significantly higher in the AD group at F8 (t=1.71, p=.037) and HFD SD was higher at left and frontal sites, including F4, Fz, F3, F7, T7, C3 and P3.

**Figure 3.**
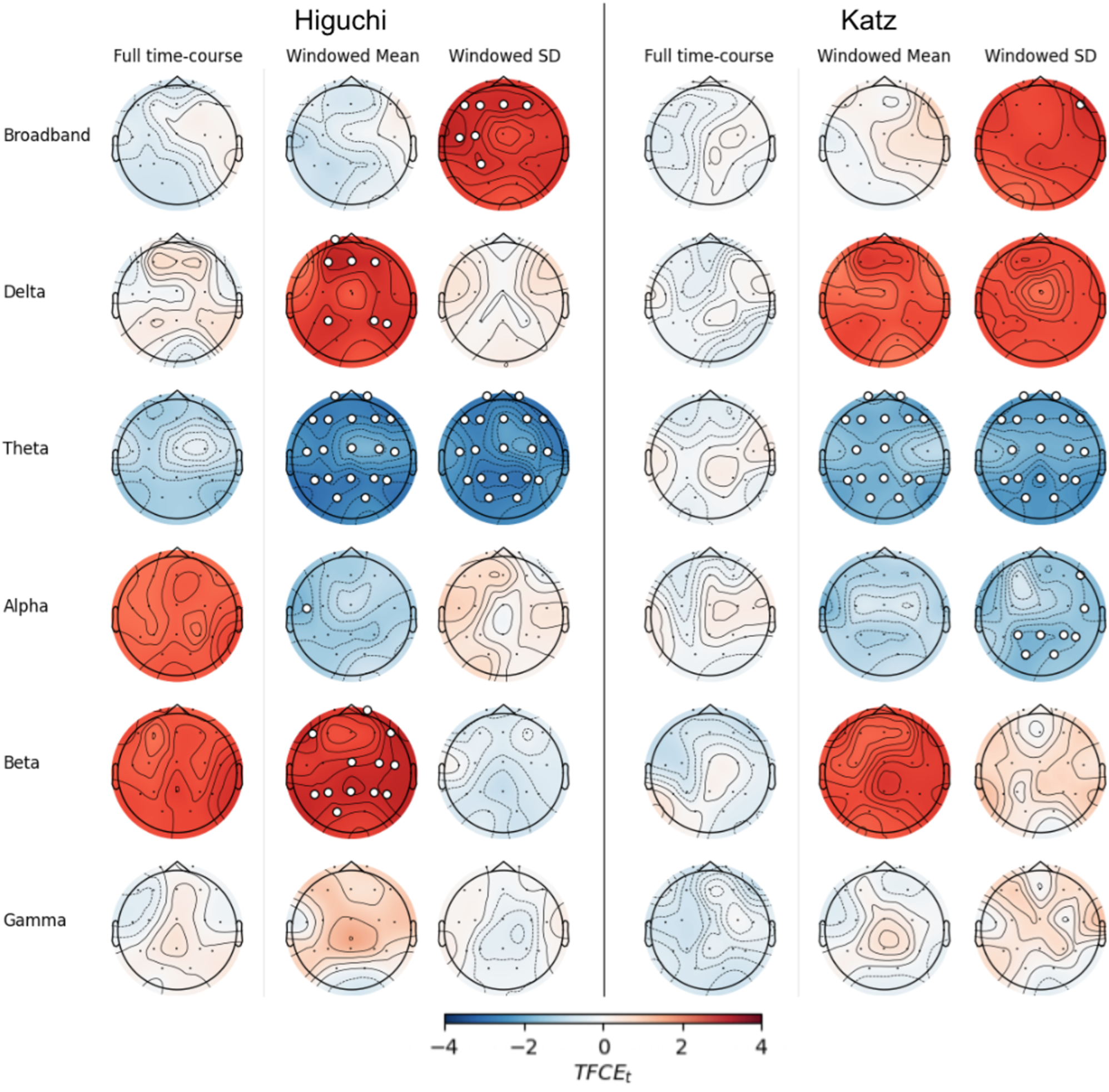
EEG fractal values that differ between AD and SCI groups. Scalp plots show differences (AD - SCI) calculated using threshold-free cluster enhancement (TFCE) for Higuchi fractal dimension (left) or Katz fractal dimension (right). Electrodes with significant differences (TFCE FWEp < .05) are marked with white circles.

The logistic regression using age to predict AD status was a significantly better fit than the intercept-only model (X^2^(1)=7.22, p=.007). Again, while full time-course HFD improved model fit relative to an age-only model, models with FDD features fit the data even better (Table 4; ΔAIC = −5.4, ΔR^2^ = .13). Furthermore, the model using Katz FDD features outperformed the model using full time-course KFD (ΔAIC = −10.7, ΔR^2^ = .42).

### 3.4. FDD is more informative for Alzheimer’s disease in most frequency bands

Next, we examined the difference between SCI and AD when FD metrics were calculated within individual frequency bands. There were no significant differences at any electrode using full time-course estimates of HFD and KFD in the delta (all *p* > .5), alpha (all *p* > .5), beta (all *p* > .1) or gamma (all *p* > .3) bands. In the theta band, no electrodes showed significant differences in full time-course HFD (all *p* > .09).

Within the delta band, mean windowed HFD was significantly higher in the AD group at frontal and parietal electrodes (all t >1.63, p < .045). The SD of windowed HFD was not significantly different between groups at any electrodes (all p > .1). Similarly, no electrodes showed significant group differences for either Katz FDD metric in the delta band (all *p* > .1). In the theta band, every electrode showed reduced Higuchi FDD measures in AD compared to SCI. Nearly all channels showed significantly lower KFD mean and SD. Finally, mean HFD was significantly higher in the AD group at central and posterior sites, as well as at Fp2, F7, and F8 (all t > 1.64, p < .049).

As with the Dem-SCI analysis, we plotted the absolute TFCE-t statistics for AD-SCI at each channel for the full time-course FD compared to TFCE-t scores obtained using FDD measures (Figure 4), as well as the difference between FDD features and the full time-course FD metrics. The most pronounced improvement was in the theta band.

**Figure 4.**
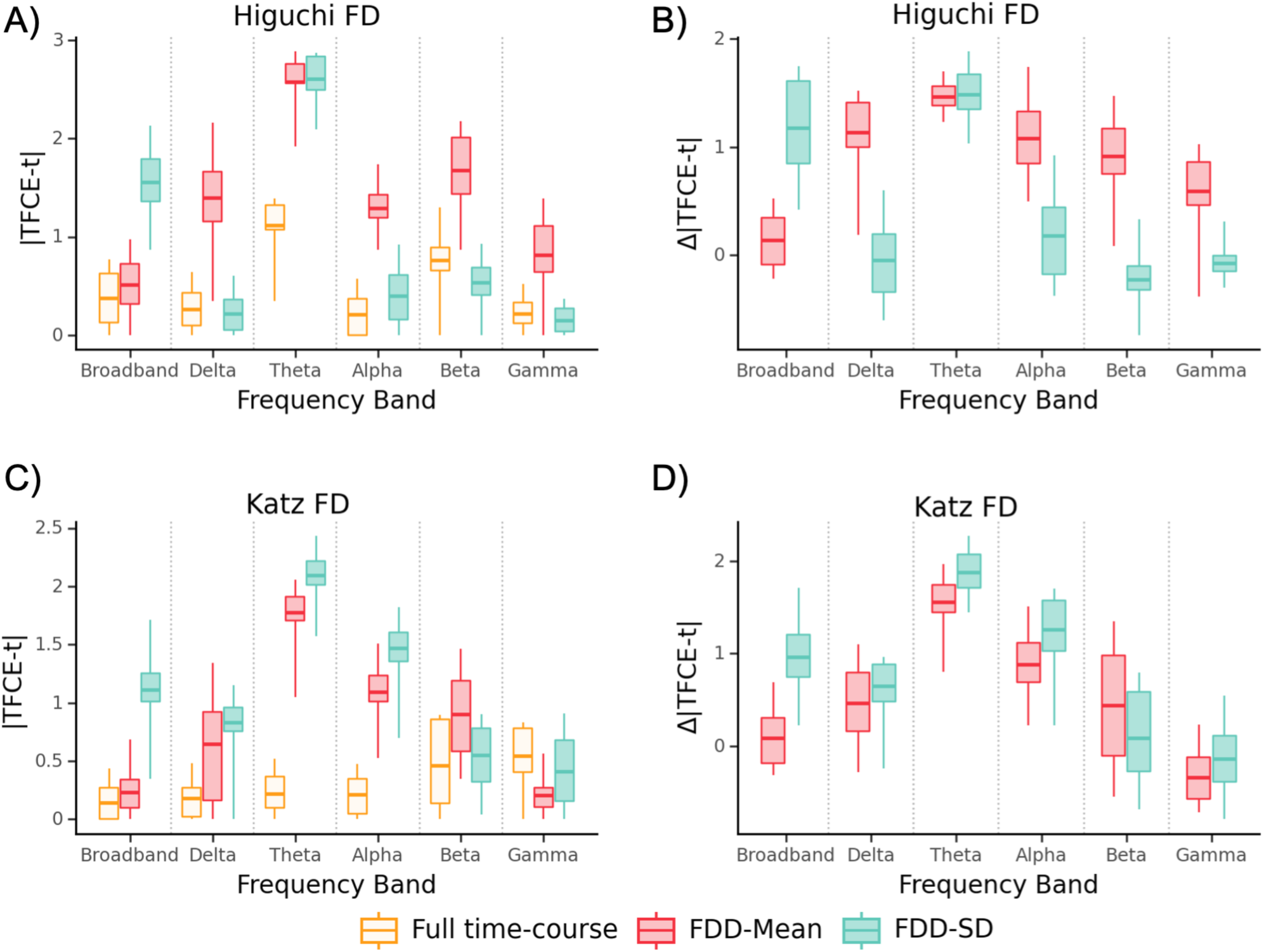
Comparison of group differences for AD-SCI with full time-course and FDD features. Average channel absolute TFCE-t statistic for AD-SCI in each frequency band using Higuchi (A) or Katz (C). Difference between FDD metrics and full time-course FD in each frequency band (B and D). Whiskers extend to minimum and maximum value.

Averaging across frequency bands, models using FDD had a lower AIC and higher R^2^ than models using full time-course estimates (ΔAIC = −2.6, ΔR^2^ = .08). Models using Higuchi FDD were nearly indistinguishable, but had improved class separation relative to full time-course models (Table 3; average ΔAIC = −0.2, ΔR^2^ = .08), while models using Katz FDD had lower AIC while showing a minimal increase in class separation. (Table 4; average ΔAIC = −2.6, ΔR^2^ = .01).

**Table 3.**
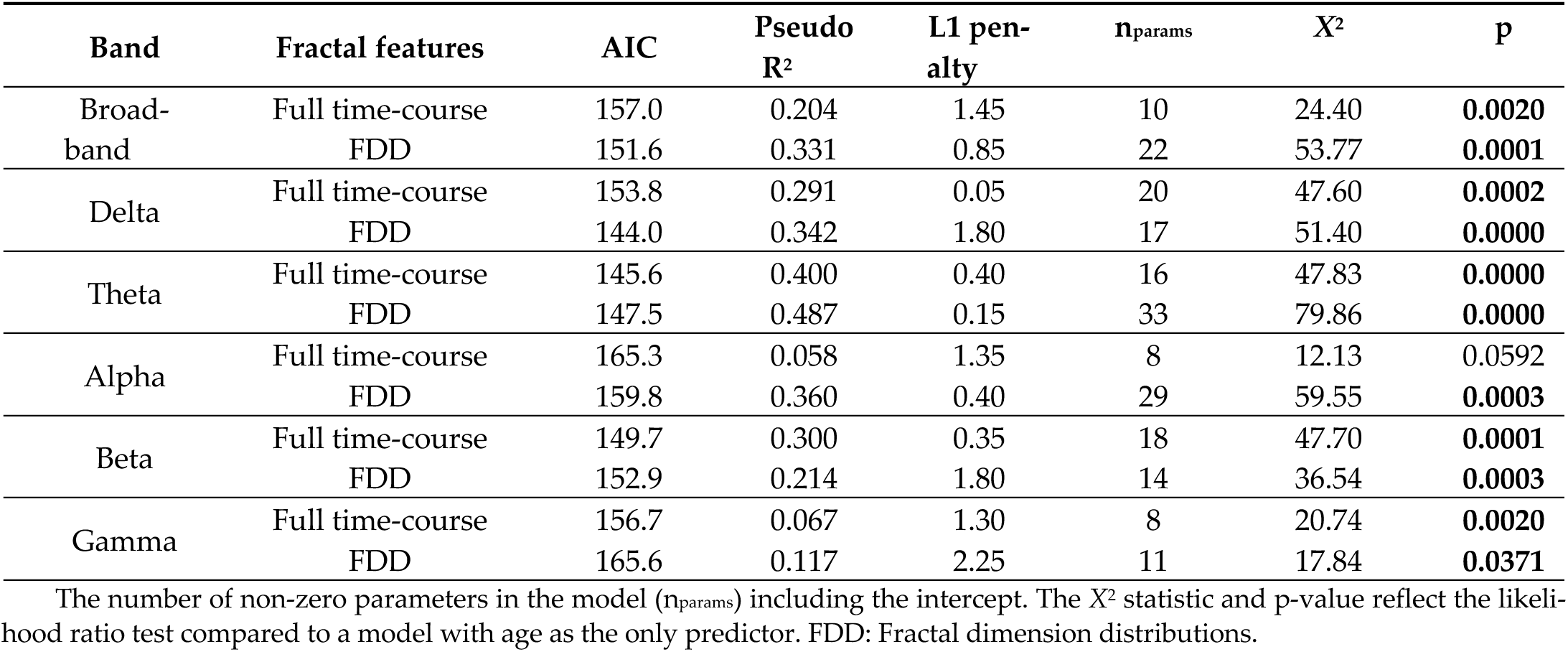
Summary of models distinguishing between AD and SCI using Higuchi fractal information.

| Band | Fractal features | AIC | Pseudo R<sup>2</sup> | L1 penalty | n<sub>params</sub> | X<sup>2</sup> | p |
| --- | --- | --- | --- | --- | --- | --- | --- |
| Broad-band | Full time-course | 157.0 | 0.204 | 1.45 | 10 | 24.40 | **0.0020** |
|  | FDD | 151.6 | 0.331 | 0.85 | 22 | 53.77 | **0.0001** |
| Delta | Full time-course | 153.8 | 0.291 | 0.05 | 20 | 47.60 | **0.0002** |
|  | FDD | 144.0 | 0.342 | 1.80 | 17 | 51.40 | **0.0000** |
| Theta | Full time-course | 145.6 | 0.400 | 0.40 | 16 | 47.83 | **0.0000** |
|  | FDD | 147.5 | 0.487 | 0.15 | 33 | 79.86 | **0.0000** |
| Alpha | Full time-course | 165.3 | 0.058 | 1.35 | 8 | 12.13 | 0.0592 |
|  | FDD | 159.8 | 0.360 | 0.40 | 29 | 59.55 | **0.0003** |
| Beta | Full time-course | 149.7 | 0.300 | 0.35 | 18 | 47.70 | **0.0001** |
|  | FDD | 152.9 | 0.214 | 1.80 | 14 | 36.54 | **0.0003** |
| Gamma | Full time-course | 156.7 | 0.067 | 1.30 | 8 | 20.74 | **0.0020** |
|  | FDD | 165.6 | 0.117 | 2.25 | 11 | 17.84 | **0.0371** |

The number of non-zero parameters in the model (n<sub>params</sub>) including the intercept. The X<sup>2</sup> statistic and p-value reflect the likelihood ratio test compared to a model with age as the only predictor. FDD: Fractal dimension distributions.

**Table 4.** Summary of models distinguishing between AD and SCI using Katz fractal information.

| Band | Fractal features | AIC | Pseudo R<sup>2</sup> | L1 penalty | n<sub>params</sub> | X<sup>2</sup> | p |
| --- | --- | --- | --- | --- | --- | --- | --- |
| Broad-band | Full time-course | 159.6 | 0.088 | 2.90 | 4 | 9.77 | **0.0076** |
|  | FDD | 148.9 | 0.507 | 0.30 | 32 | 76.48 | **0.0000** |
| Delta | Full time-course | 167.6 | 0.009 | 2.45 | 5 | 3.83 | 0.2804 |
|  | FDD | 155.5 | 0.224 | 2.15 | 15 | 35.90 | **0.0006** |
| Theta | Full time-course | 156.8 | 0.341 | 0.15 | 19 | 42.64 | **0.0005** |
|  | FDD | 154.8 | 0.126 | 2.25 | 8 | 22.64 | **0.0009** |
| Alpha | Full time-course | 162.7 | 0.165 | 1.25 | 13 | 24.68 | **0.0101** |
|  | FDD | 164.9 | 0.107 | 2.75 | 8 | 12.49 | 0.0519 |
| Beta | Full time-course | 159.1 | 0.252 | 0.65 | 15 | 32.26 | **0.0022** |
|  | FDD | 163.6 | 0.049 | 4.45 | 5 | 7.83 | **0.0496** |
| Gamma | Full time-course | 168.5 | 0.000 | 3.80 | 4 | 0.93 | 0.6266 |
|  | FDD | 161.7 | 0.233 | 1.20 | 21 | 41.69 | **0.0020** |

The number of non-zero parameters in the model (n<sub>params</sub>) including the intercept. The X<sup>2</sup> statistic and p-value reflect the likelihood ratio test compared to a model with age as the only predictor. FDD: Fractal dimension distributions.

Models using Higuchi FDD outperformed models using full time-course HFD in the delta (Table 3; ΔAIC = −9.8, ΔR^2^ = .05) and alpha bands (ΔAIC = −5.4, ΔR^2^ = .30). In the theta and gamma bands, the windowed model separated the classes better, but had worse expected prediction error (ΔAIC = 2.0, ΔR^2^ = .09; ΔAIC = 8.9, ΔR^2^ = .05). The model using FDD features had lower performance than the models using full time-course HFD in the beta band (ΔAIC = 3.2, ΔR^2^ = -.09).

The models using KFD were similarly mixed. Models using FDD features outperformed models using full time-course FD in the delta (ΔAIC = −12.1, ΔR^2^ = .22) and gamma bands (ΔAIC = −6.8, ΔR^2^ = 0.23), but underperformed models using full time-course FD in the alpha (ΔAIC = 2.2, ΔR^2^ = -.06) and beta bands (ΔAIC = 4.4, ΔR^2^ = -.20). In the theta band, models based on FDD had lower AIC but also lower class separation (ΔAIC = −2.0, ΔR^2^ = -.22).

Across all models comparing AD and SCI, the models with the lowest AIC and largest R^2^ used Higuchi FDD in the delta band (AIC=144.0, R^2^ = 0.342, X^2^(17)=51.40, p < .001) or Katz FDD with broadband EEG (AIC=148.9, R^2^ = 0.507, X^2^(32)=76.48, p < .001).

### 3.5. Features useful for distinguishing AD and dementia partially overlap

Are the features that are useful for distinguishing between SCI and any dementia the same as the features that are useful for distinguishing between SCI and AD-dementia? To answer this question, we examined the regression coefficients from each of the final models. In these LASSO regressions, L1 regularization selects variables that relate to the dependent variable, and sets all other coefficients to zero. We examined which channels, frequencies, and FD analysis methods were retained in these models as non-zero coefficients. As shown in Figure 5, while the regularized logistic regression indicated that some features were useful for both models, others features were only useful in one of the models but not the other. Across the fractal variability models, Dementia-SCI models used a total of 122 EEG features while AD-SCI models used 191, with 89 features appearing in both models.

**Figure 5.**
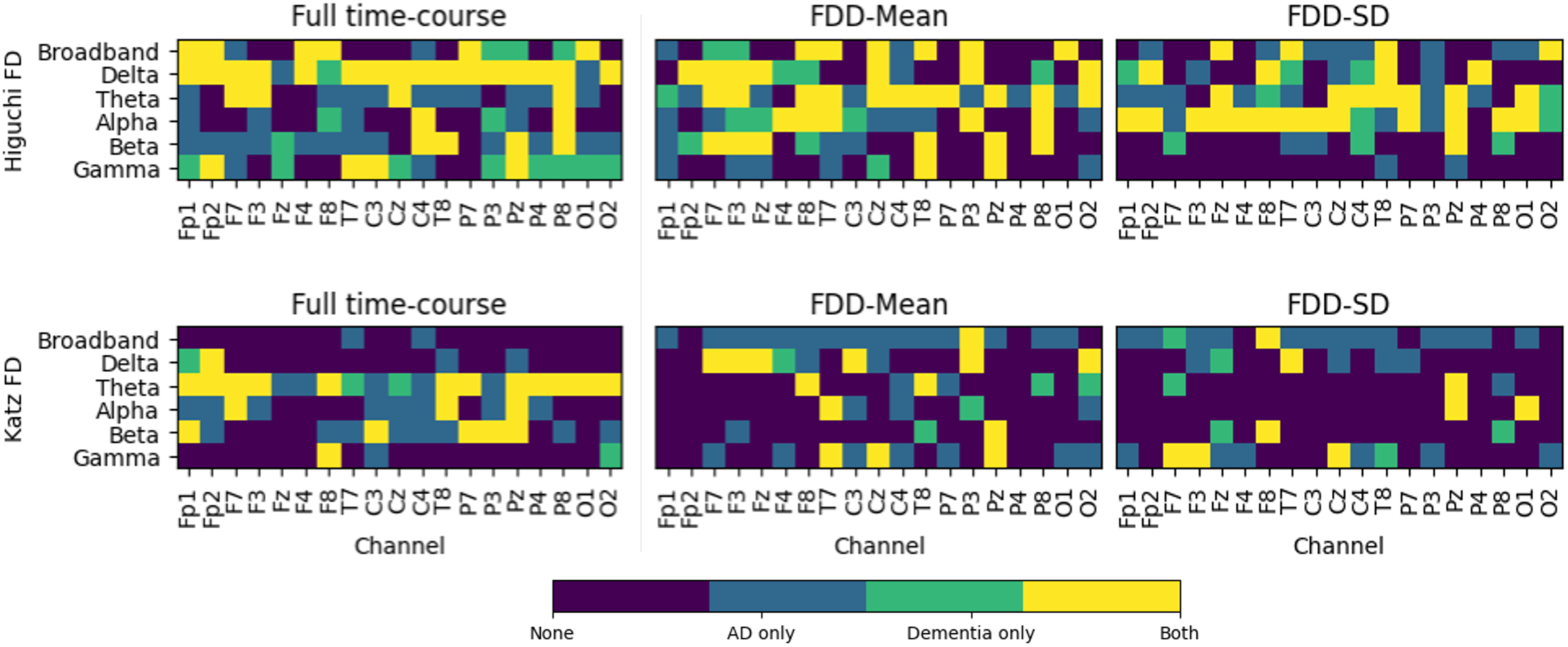
Channels used in final models. Channels with non-zero coefficients in the regularized logistic regression using full time-course FD (left) or FDD (center and right) features calculated using Higuchi (top) or Katz (bottom) methods.

### 3.6. FDD provides information about dementia across different window lengths

To understand whether our results depended on the length of the window used to calculate FDD features, we repeated the analysis using three other window sizes — 0.5 s, 5 s, 10 s — each with a 50% overlap. We estimated ideal *k*_max_ = 25 for 0.5 s and 5 s windows, and *k*_max_ = 28 for 10 s windows. Models using FDD continued to generally outperform models using full time-course FD values, with better performance in broadband for both Dementia-SCI and AD-SCI. (see Supplemental Tables 1-4). Across analyses comparing Dementia-SCI within specific frequency bands, models using FDD features calculated with 1 s windows outperformed models using full time-course FD in 8/10 cases. Similarly, using 0.5 s or 5 s windows resulted in better performance with FDD features in 9/10 cases and FDD calculated from 10 s windows produced better performance in 6/10 cases. When modeling AD-SCI, FDD with 1 s windows outperformed full time-course FD in 5/10 models. Changing the window size resulted in similar performance for 0.5 s (5/10 models), 5 s (4/10 models), or 10 s (6/10) durations.

## 4. Discussion

In this study, we present the FDD metric, a novel method for quantifying the distribution of fractal dimension values over time. We then apply FDD to examine signal complexity in Alzheimer’s disease. We show that FDD carries information above and beyond the full time-course fractal dimension of a signal, and that FDD features are useful at distinguishing individuals diagnosed with dementia from individuals with SCI, as well as individuals with AD-dementia from individuals with SCI. Using both Higuchi and Katz algorithms, FDD calculated from broadband EEG revealed more significant differences between groups than full time-course fractal values (Figures 1 and 2). Models using FDD features were also better (as measured by lower AIC and higher R^2^) than models using full time-course FD (Tables 1 and 2). FDD features also demonstrated more group differences, larger effect sizes, and better models across most frequency bands (Tables 1 and 2; Figures 1 and 2).

When comparing SCI to dementia or SCI to AD-dementia, the most widespread group differences were observed in the theta band (Figures 1 and 3). Theta band activity has long been associated with memory performance[38]. This is not entirely surprising, since memory loss is a hallmark of dementia. However, there are many open questions related to theta oscillations in human neocortex, such as whether observed associations between memory function and theta power arise from focal changes in theta power or rather as an overall shift in the power spectrum[39].

A second goal of this work was to understand whether the distribution of fractal dimension scores could also elucidate characteristics of AD-dementia. FDD features again revealed more electrodes with significant AD-SCI differences than full time-course FD, both in broadband and the majority of traditional frequency bands (Figures 3 and 4). Using broadband data, FDD features produced better models for both HFD and KFD (Tables 3 and 4). Models using FDD to distinguish between AD and SCI were better in the delta band and slightly better on average.

Previous work has found lower HFD in AD[19–22]. Similarly, prior studies have examined FD separately within canonical frequency bands, and find lower KFD and HFD in AD[11,26,40]. Using FDD, we replicate this decrease in FD for the dementia and AD-dementia within the theta and alpha bands. Using full time-course FD, we find non-significant trends toward decreases in FD using full time-course broadband HFD and KFD. Why do we find a non-significant trend where other studies showed a significant decrease? We compare AD/Dementia to SCI, whereas other work used healthy older adults recorded in a laboratory setting[11,18]. There may also be an important difference between FD calculated from EEG compared to MEG[19,20]. Moreover, as discussed above (see 2.3), computing HFD relies on the k_max_ parameter, and previous investigations of HFD in dementia did not calculate k_max_ using the approach based on time-course length[34]. Our proposed FDD metric did reliably identify reduced complexity, for both HFD and KFD.

Our results also highlight the importance of distinguishing between dementia caused by AD from non-AD dementia. Particularly when using FDD, the Dementia and SCI groups demonstrate significant differences across nearly the entire scalp (Figure 1). In contrast, significant differences between AD and SCI were restricted to fewer electrodes, and appeared primarily in the theta band (Figures 3 and 4). Moreover, while 73% of the FDD features that were retained in the Dementia-SCI models were also retained in AD-SCI models (Figure 5), less than half of the features used in the AD-SCI models appeared in the Dementia-SCI models (47%). In other words, while some EEG signals are generally useful for detecting cognitive impairment, our results suggest that EEG is also sensitive to additional signals which are uniquely important for detecting AD. Whether FDD is useful for distinguishing between AD and non-AD dementia cases will require additional evidence with larger populations of patients with dementia.

AD is a neurodegenerative disease with a distinct progression of physical and cognitive symptoms. Its pathology is characterized by two features: extracellular deposits of beta-amyloid plaques, and intracellular accumulations of abnormally phosphorylated tau, called neurofibrillary tangles[41]. Patients with Alzheimer’s dementia exhibit widespread amyloid plaques and neurofibrillary tangles throughout the brain, but beta-amyloid and tau start to amass long before severe cognitive symptoms appear[42]. One origin of reduced EEG complexity in AD patients may be an abnormal cortical excitation/inhibition ratio[43]; beta-amyloid is linked to neuronal hypoactivity, and tau is associated with neuronal hyperactivity[44]. Decreases in EEG complexity might also arise from general neurodegeneration, with fewer neurons and fewer interactions between neurons[45]. However, more work is needed to clarify any associations between beta-amyloid and tau abnormalities, cognitive decline, and EEG complexity.

Researchers investigating FD in EEG might also benefit from examining FDD features as a complement to full time-course FD methods. Previous work has used FD in EEG signals to identify a variety of neuropsychological and neurocognitive conditions. EEG complexity and FD in schizophrenia has been widely investigated, with studies reporting both increased and decreased FD based on symptomatology, age, and medication status [46–49]. In schizophrenia, the FD also has strong predictive power, and it has been used to distinguish individuals with schizophrenia from healthy controls with high accuracy [50]. Other work has examined FD in mood and cognitive disorders, with increased FD reported in depression [46,51,52], bipolar disorder [53], and attention deficit hyperactivity disorder [54].

Our introduction of FDD is focused on the mean and standard deviation of FD compute across multiple moving windows. However, future work could investigate other methods of assessing changes in fractal content over time. For instance, higher-order distributional summary statistics, such as skewness or kurtosis could be included. Similarly, time-sensitive measures, such as autoregressive variance might provide additional unique information that could be used to identify or distinguish neurodegenerative conditions.

One additional potential advantage of FDD over full time-course fractal measures is that it allows for momentary artifacts to be excluded from the data. If artifacts contaminate a portion of an EEG recording, that artifact could bias estimates of FD. A windowed approach like FDD makes it trivially easy to handle events such as jump artifacts or movement artifacts — simply exclude windows with artifacts from the analysis. Thus, FDD offers the potential to recover usable EEG recordings from a broader range of patients.

While our results suggest that models trained using FDD could be a useful early screening tool in clinical settings, future studies will be required to determine optimal FDD parameters. We expect that FDD will be most useful in a clinical setting when used alongside other EEG features, such as spectral power, or combined with molecular or liquid biomarkers.

The current manuscript has several limitations. The dataset contains only 13 patients with non-AD dementia. This means we were not well-positioned to directly address how well FDD might distinguish between different dementia subtypes. Moreover, our data are cross-sectional. Longitudinal studies will be needed to assess the prospective utility of FDD. Our goal was to introduce FDD, rather than propose exact parameters values. Thus, we did not exhaustively test potential values for parameters, such as window length.

## 5. Conclusions

We propose a novel method, FDD, to investigate fractal dimensions within and across frequency bands in restingstate EEG data. Our results extend previous work linking differences in EEG spectral content and fractal dimension to AD and non-AD dementia. In broadband EEG, and within most of the traditional frequency bands, FDD revealed stronger group differences and more informative features than full time-course FD for both dementia and AD-dementia. Moreover, regularized linear regressions using FDD features were better at accounting for differences between unimpaired subjects and subjects with dementia than models using full time-course fractal dimension. Overall, our findings demonstrate that FDD can provide information about cognitive status and AD diagnosis from resting-state EEG, above and beyond traditional full time-course methods.

## Author Contributions

Conceptualization, K.J.Y, C.Q, and C.L.; methodology, K.J.Y.; software, K.J.Y., G.B., and C.Q..; formal analysis, K.J.Y.; resources, D.A.M. and S.G.; data curation, C.Q.; writing—original draft preparation, K.J.Y.; writing—review and editing, K.J.Y., G.B., R.G., D.A.M, C.Q., and C.L.; visualization, K.J.Y.; supervision, D.A.M. and S.G.; funding acquisition, S.G. and C.L. All authors have read and agreed to the published version of the manuscript.

## Funding

This work was supported by SPARK Neuro Inc. The funder was not involved in the study design, collection, analysis, interpretation of data, the writing of the article, or the decision to submit it for publication.

## Institutional Review Board Statement

This study was conducted in accordance with the Declaration of Helsinki. The Institutional Review Board of Saint John’s Cancer Institute gave ethical approval for this work (Protocol JWCI-19-1101).

## Informed Consent Statement

Informed consent was obtained from all subjects involved in the study.

## Data Availability Statement

Restrictions apply to the availability of these data. Data were obtained from the Pacific Neuroscience Institute [55] and can be obtained from D.A.M with permission of D.A.M.

## Conflicts of Interest

K.J.Y, G.B, S.G, C.Q., and C.L were employed by SPARK Neuro Inc., a medical technology company developing diagnostic aids to help clinicians identify and assess neurodegenerative diseases, during this study. Remaining authors have no competing interests to declare.

