## Supplementary Materials for "Fractal dimension distributions of resting-state EEG improve detection of dementia and Alzheimer’s disease compared to traditional fractal analysis"

**Estimating $k_{max}$ for Higuchi's fractal dimension**

Wanliss & Wanliss performed a series of simulations using fractional Brownian motion [34]. For each, they calculated Higuchi fractal dimension (HFD) across a range of $k_{max}$ values and compared the computed HFD to the expected theoretical fractal dimension of the signal. They then took the geometric mean of these curves and used a sum of sines equation to estimate the best fit line. This produced the following formula for estimating the optimal $k_{max}$ for $N$ samples:

$$k_{max} = [A_1 \sin(B_1 * N + C_1) + A_2 \sin(B_2 * N + C_2)] \tag{4}$$

Here, [ ] means only the integer component of the $k_{max}$ is used. Estimates for the best fitting equation are given in Supplemental Table 1.

**Table S1. Estimates of best fitting parameters for $k_{max}$ function**

| Parameter | Estimate |
|---|---|
| $A_1$ | 129.8 ± 3.0 |
| $B_1$ | (1.292 ± 0.045) x $10^{-5}$ |
| $C_1$ | 0.04488 ± 0.0255 |
| $A_2$ | 18.82 ± 2.56 |
| $B_2$ | (6.488 ± 0.280) x $10^{-5}$ |
| $C_2$ | 1.332 ± 0.220 |

We assumed a Gaussian distribution of parameters, then sampled from these distributions and generated 10,000 $k_{max}$ values and selected the modal response. This approach resulted in $k_{max}$ = 108 for calculating HFD with the full time-course, and $k_{max}$ = 25 for calculating HFD for the windowed data.

**Figure S1. Channel correlations for full time-course fractal dimension**. Heatmap showing channel-by-channel correlations for HFD (upper right triangle) and KFD (lower left triangle). Note that all resulting correlations are positive.

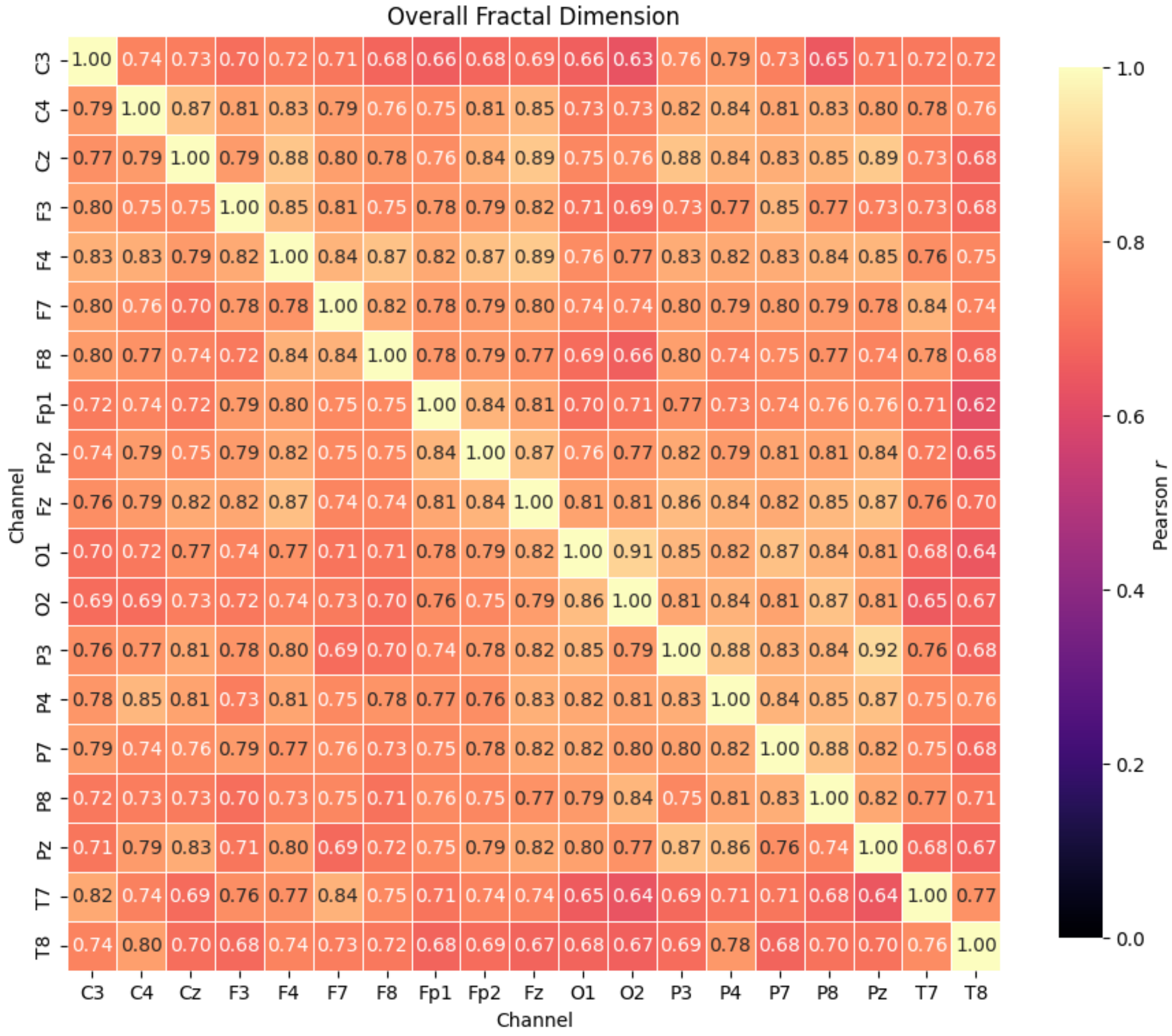

**Figure S2. Channel correlations for windowed fractal mean**. Heatmap showing channel-by-channel correlations for HFD (upper right triangle) and KFD (lower left triangle). Note that all resulting correlations are positive.

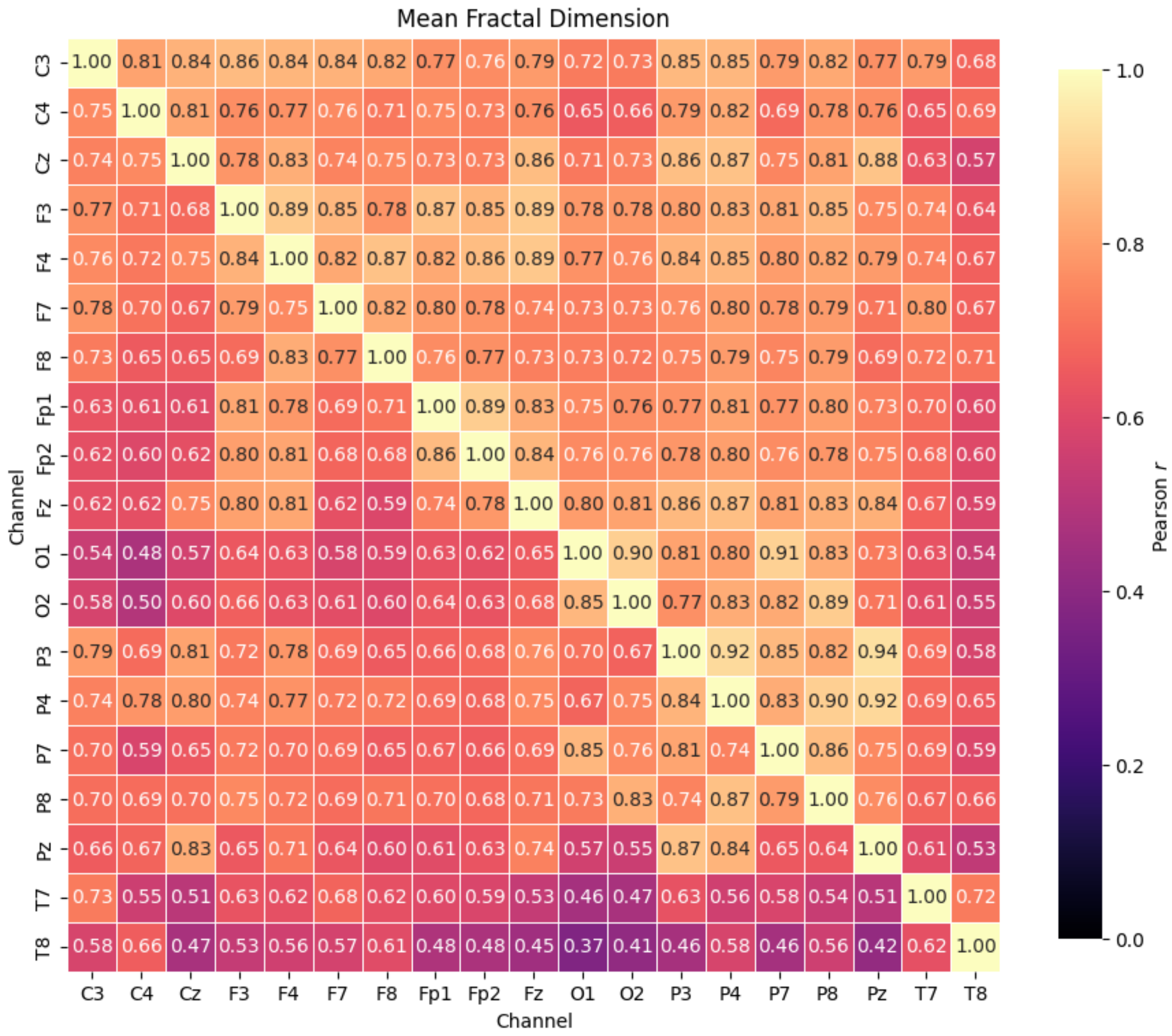

**Figure S3. Channel correlations for windowed fractal standard deviation**. Heatmap showing channel-by-channel correlations for HFD (upper right triangle) and KFD (lower left triangle). Note that all resulting correlations are positive.

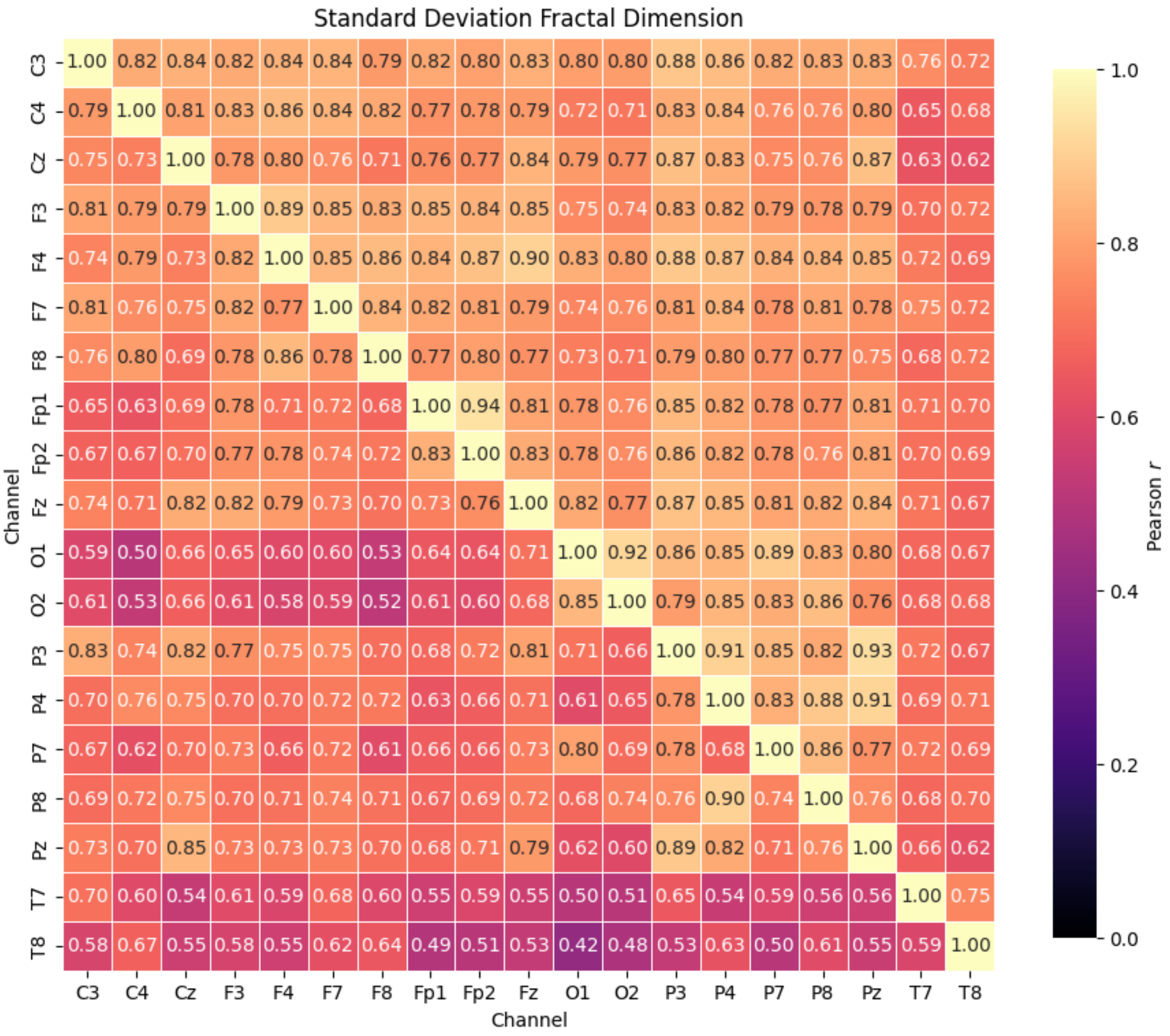

**Table S2. Summary of models predicting Dementia vs SCI using Higuchi fractal information with different window lengths**

| Band | Model | Window | AIC | Pseudo $R^2$ | L1 penalty | $n_{params}$ | $X2$ | p |
|---|---|---|---|---|---|---|---|---|
| Broadband | Full time-course | | 180.3 | 0.290 | 1.60 | 11 | 24.74 | **0.0033** |
| | FDD features | 0.5 s | 171.6 | 0.310 | 3.75 | 5 | 21.41 | **0.0001** |
| | FDD features | 1 s | 163.6 | 0.446 | 1.85 | 6 | 47.40 | **0.0111** |
| | FDD features | 5 s | 171.7 | 0.339 | 4.10 | 6 | 23.31 | **0.0001** |
| | FDD features | 10 s | 173.0 | 0.339 | 3.40 | 7 | 24.07 | **0.0002** |
| Delta | Full time-course | | 178.8 | 0.425 | 0.25 | 19 | 42.20 | **0.0006** |
| | FDD features | 0.5 s | 182.3 | 0.252 | 0.65 | 11 | 22.70 | **0.0069** |
| | FDD features | 1 s | 176.1 | 0.397 | 1.90 | 19 | 44.91 | **0.0003** |
| | FDD features | 5 s | 178.7 | 0.371 | 3.00 | 10 | 24.32 | **0.0020** |
| | FDD features | 10 s | 180.9 | 0.409 | 1.75 | 13 | 28.17 | **0.0030** |
| Theta | Full time-course | | 174.9 | 0.269 | 1.70 | 6 | 20.13 | **0.0005** |
| | FDD features | 0.5 s | 164.1 | 0.582 | 0.10 | 34 | 86.91 | **0.0000** |
| | FDD features | 1 s | 159.6 | 0.592 | 0.85 | 23 | 69.44 | **0.0000** |
| | FDD features | 5 s | 165.4 | 0.494 | 2.25 | 9 | 35.61 | **0.0000** |
| | FDD features | 10 s | 176.3 | 0.259 | 3.25 | 6 | 18.79 | **0.0009** |
| Alpha | Full time-course | | 182.0 | 0.262 | 1.85 | 6 | 13.04 | **0.0111** |
| | FDD features | 0.5 s | 171.0 | 0.467 | 0.65 | 20 | 52.08 | **0.0000** |
| | FDD features | 1 s | 166.6 | 0.644 | 0.75 | 25 | 66.42 | **0.0000** |
| | FDD features | 5 s | 170.0 | 0.476 | 0.90 | 23 | 59.06 | **0.0000** |
| | FDD features | 10 s | 173.8 | 0.507 | 0.95 | 21 | 51.25 | **0.0001** |
| Beta | Full time-course | | 179.5 | 0.240 | 2.15 | 7 | 17.57 | **0.0035** |
| | FDD features | 0.5 s | 171.0 | 0.349 | 1.05 | 12 | 36.06 | **0.0001** |
| | FDD features | 1 s | 173.0 | 0.377 | 1.90 | 14 | 38.05 | **0.0002** |
| | FDD features | 5 s | 169.2 | 0.407 | 1.00 | 20 | 53.84 | **0.0000** |
| | FDD features | 10 s | 175.4 | 0.565 | 0.45 | 28 | 63.63 | **0.0001** |
| Gamma | Full time-course | | 184.4 | 0.298 | 0.60 | 14 | 26.65 | **0.0087** |
| | FDD features | 0.5 s | 176.3 | 0.389 | 0.25 | 16 | 38.77 | **0.0004** |
| | FDD features | 1 s | 184.7 | 0.220 | 4.75 | 5 | 8.33 | **0.0396** |
| | FDD features | 5 s | 182.3 | 0.279 | 1.60 | 12 | 24.77 | **0.0058** |
| | FDD features | 10 s | 187.2 | 0.096 | 4.10 | 3 | 1.85 | 0.1742 |

The number of non-zero parameters in the model ($n_{params}$) including the intercept.

**Table S3. Summary of models predicting Dementia vs SCI using Katz fractal information with different window lengths**

| Band | Model | Window | AIC | Pseudo $R^2$ | L1 penalty | $n_{params}$ | *X*2 | p |
|---|---|---|---|---|---|---|---|---|
| Broadband | Full time-course | | 189.2 | 0.058 | 5.65 | 2 | -2.17 | 1.0000 |
| | FDD features | 0.5 s | 175.6 | 0.269 | 4.65 | 6 | 19.48 | **0.0006** |
| | FDD features | 1 s | 176.5 | 0.299 | 0.25 | 19 | 16.54 | **0.0006** |
| | FDD features | 5 s | 182.0 | 0.152 | 4.90 | 5 | 11.05 | **0.0115** |
| | FDD features | 10 s | 185.4 | 0.153 | 4.15 | 6 | 9.64 | **0.0470** |
| Delta | Full time-course | | 190.8 | 0.087 | 3.70 | 4 | 0.28 | 0.8698 |
| | FDD features | 0.5 s | 184.0 | 0.476 | 1.25 | 24 | 47.09 | **0.0014** |
| | FDD features | 1 s | 183.0 | 0.261 | 3.55 | 11 | 22.09 | **0.0086** |
| | FDD features | 5 s | 187.3 | 0.486 | 0.70 | 28 | 51.76 | **0.0019** |
| | FDD features | 10 s | 189.6 | 0.202 | 4.75 | 6 | 5.45 | 0.2440 |
| Theta | Full time-course | | 179.6 | 0.398 | 0.60 | 16 | 35.46 | **0.0013** |
| | FDD features | 0.5 s | 165.0 | 0.592 | 0.45 | 28 | 74.03 | **0.0000** |
| | FDD features | 1 s | 174.8 | 0.358 | 2.40 | 8 | 24.20 | **0.0005** |
| | FDD features | 5 s | 167.8 | 0.653 | 0.85 | 26 | 67.27 | **0.0000** |
| | FDD features | 10 s | 175.6 | 0.603 | 0.80 | 27 | 61.45 | **0.0001** |
| Alpha | Full time-course | | 187.0 | 0.223 | 3.50 | 5 | 6.03 | 0.1100 |
| | FDD features | 0.5 s | 185.0 | 0.211 | 5.40 | 5 | 8.04 | **0.0451** |
| | FDD features | 1 s | 186.1 | 0.229 | 5.05 | 6 | 8.96 | 0.0622 |
| | FDD features | 5 s | 190.2 | 0.125 | 4.50 | 5 | 2.85 | 0.4160 |
| | FDD features | 10 s | 190.6 | 0.058 | 5.10 | 3 | -1.52 | 1.0000 |
| Beta | Full time-course | | 188.1 | 0.133 | 2.45 | 7 | 8.93 | 0.1120 |
| | FDD features | 0.5 s | 186.6 | 0.280 | 3.05 | 11 | 18.40 | **0.0308** |
| | FDD features | 1 s | 178.9 | 0.270 | 4.25 | 7 | 18.12 | **0.0028** |
| | FDD features | 5 s | 177.0 | 0.429 | 2.65 | 12 | 30.05 | **0.0008** |
| | FDD features | 10 s | 182.0 | 0.302 | 2.80 | 11 | 23.00 | **0.0062** |
| Gamma | Full time-course | | 189.1 | 0.135 | 2.85 | 4 | 1.99 | 0.3705 |
| | FDD features | 0.5 s | 186.9 | 0.329 | 2.40 | 12 | 20.17 | **0.0277** |
| | FDD features | 1 s | 194.1 | 0.232 | 4.70 | 9 | 6.98 | 0.4313 |
| | FDD features | 5 s | 185.9 | 0.145 | 4.65 | 4 | 5.13 | 0.0768 |
| | FDD features | 10 s | 189.0 | 0.036 | 4.35 | 4 | 2.04 | 0.3605 |

The number of non-zero parameters in the model ($n_{params}$) including the intercept.

**Table S4. Summary of models predicting AD vs SCI using Katz fractal information with different window lengths**

| Band | Model | Window | AIC | Pseudo $R^2$ | L1 penalty | $n_{params}$ | *X2* | p |
|---|---|---|---|---|---|---|---|---|
| Broadband | Full time-course | | 157.0 | 0.204 | 1.45 | 10 | 24.40 | **0.0020** |
| | FDD features | 0.5 s | 151.1 | 0.341 | 0.60 | 25 | 60.24 | **0.0000** |
| | FDD features | 1 s | 151.6 | 0.331 | 0.85 | 22 | 53.77 | **0.0001** |
| | FDD features | 5 s | 152.5 | 0.380 | 0.55 | 27 | 62.88 | **0.0000** |
| | FDD features | 10 s | 150.2 | 0.292 | 0.90 | 21 | 53.23 | **0.0000** |
| Delta | Full time-course | | 153.8 | 0.291 | 0.05 | 20 | 47.60 | **0.0002** |
| | FDD features | 0.5 s | 155.1 | 0.253 | 0.25 | 18 | 42.30 | **0.0004** |
| | FDD features | 1 s | 144.0 | 0.342 | 1.80 | 17 | 51.40 | **0.0000** |
| | FDD features | 5 s | 156.4 | 0.254 | 2.60 | 12 | 28.98 | **0.0013** |
| | FDD features | 10 s | 159.9 | 0.117 | 3.00 | 9 | 19.47 | **0.0068** |
| Theta | Full time-course | | 145.6 | 0.400 | 0.40 | 16 | 47.83 | **0.0000** |
| | FDD features | 0.5 s | 155.6 | 0.185 | 1.60 | 9 | 23.82 | **0.0012** |
| | FDD features | 1 s | 147.5 | 0.487 | 0.15 | 33 | 79.86 | **0.0000** |
| | FDD features | 5 s | 154.6 | 0.205 | 2.50 | 9 | 24.84 | **0.0008** |
| | FDD features | 10 s | 159.8 | 0.068 | 3.45 | 6 | 13.57 | **0.0088** |
| Alpha | Full time-course | | 165.3 | 0.058 | 1.35 | 8 | 12.13 | 0.0592 |
| | FDD features | 0.5 s | 160.4 | 0.156 | 1.15 | 11 | 22.94 | **0.0063** |
| | FDD features | 1 s | 159.8 | 0.360 | 0.40 | 29 | 59.55 | **0.0003** |
| | FDD features | 5 s | 165.7 | 0.039 | 4.15 | 5 | 5.69 | 0.1279 |
| | FDD features | 10 s | 158.6 | 0.439 | 0.25 | 31 | 64.77 | **0.0002** |
| Beta | Full time-course | | 149.7 | 0.300 | 0.35 | 18 | 47.70 | **0.0001** |
| | FDD features | 0.5 s | 160.5 | 0.459 | 0.15 | 34 | 68.92 | **0.0002** |
| | FDD features | 1 s | 152.9 | 0.214 | 1.80 | 14 | 36.54 | **0.0003** |
| | FDD features | 5 s | 149.7 | 0.547 | 0.10 | 37 | 85.73 | **0.0000** |
| | FDD features | 10 s | 166.0 | 0.059 | 3.85 | 6 | 7.42 | 0.1155 |
| Gamma | Full time-course | | 156.7 | 0.067 | 1.30 | 8 | 20.74 | **0.0020** |
| | FDD features | 0.5 s | 157.0 | 0.107 | 0.55 | 10 | 24.39 | **0.0020** |
| | FDD features | 1 s | 165.6 | 0.117 | 2.25 | 11 | 17.84 | **0.0371** |
| | FDD features | 5 s | 164.7 | 0.175 | 1.55 | 14 | 24.73 | **0.0161** |
| | FDD features | 10 s | 166.7 | 0.020 | 3.60 | 6 | 6.68 | 0.1537 |

The number of non-zero parameters in the model ($n_{params}$) including the intercept.

**Table S5. Summary of models predicting AD vs SCI using Katz fractal information with different window lengths**

| Band | Model | Window | AIC | Pseudo $R^2$ | L1 penalty | $n_{params}$ | *X*2 | p |
|---|---|---|---|---|---|---|---|---|
| Broadband | Full time-course | | 159.6 | 0.088 | 2.90 | 4 | 9.77 | **0.0076** |
| | FDD features | 0.5 s | 155.1 | 0.301 | 1.10 | 22 | 50.29 | **0.0002** |
| | FDD features | 1 s | 148.9 | 0.507 | 0.30 | 32 | 76.48 | **0.0000** |
| | FDD features | 5 s | 156.2 | 0.213 | 1.10 | 18 | 41.15 | **0.0005** |
| | FDD features | 10 s | 158.1 | 0.322 | 0.75 | 22 | 47.24 | **0.0005** |
| Delta | Full time-course | | 167.6 | 0.009 | 2.45 | 5 | 3.83 | 0.2804 |
| | FDD features | 0.5 s | 150.6 | 0.262 | 1.95 | 14 | 38.82 | **0.0001** |
| | FDD features | 1 s | 155.5 | 0.224 | 2.15 | 15 | 35.90 | **0.0006** |
| | FDD features | 5 s | 167.9 | 0.117 | 2.80 | 14 | 21.49 | **0.0436** |
| | FDD features | 10 s | 162.3 | 0.283 | 1.6 | 20 | 39.14 | **0.0027** |
| Theta | Full time-course | | 156.8 | 0.341 | 0.15 | 19 | 42.64 | **0.0005** |
| | FDD features | 0.5 s | 154.4 | 0.234 | 1.50 | 14 | 35.00 | **0.0005** |
| | FDD features | 1 s | 154.8 | 0.126 | 2.25 | 8 | 22.64 | **0.0009** |
| | FDD features | 5 s | 150.9 | 0.215 | 2.40 | 10 | 30.49 | **0.0002** |
| | FDD features | 10 s | 151.5 | 0.263 | 1.85 | 12 | 33.88 | **0.0002** |
| Alpha | Full time-course | | 162.7 | 0.165 | 1.25 | 13 | 24.68 | **0.0101** |
| | FDD features | 0.5 s | 162.6 | 0.020 | 5.25 | 3 | 4.78 | **0.0289** |
| | FDD features | 1 s | 164.9 | 0.107 | 2.75 | 8 | 12.49 | 0.0519 |
| | FDD features | 5 s | 171.0 | 0.020 | 2.65 | 10 | 10.35 | 0.2410 |
| | FDD features | 10 s | 166.2 | 0.000 | 3.40 | 5 | 5.18 | 0.1589 |
| Beta | Full time-course | | 159.1 | 0.252 | 0.65 | 15 | 32.26 | **0.0022** |
| | FDD features | 0.5 s | 161.9 | 0.429 | 0.15 | 33 | 65.54 | **0.0003** |
| | FDD features | 1 s | 163.6 | 0.049 | 4.45 | 5 | 7.83 | **0.0496** |
| | FDD features | 5 s | 148.8 | 0.330 | 0.85 | 26 | 64.59 | **0.0000** |
| | FDD features | 10 s | 145.8 | 0.272 | 2.05 | 14 | 43.59 | **0.0000** |
| Gamma | Full time-course | | 168.5 | 0.000 | 3.80 | 4 | 0.93 | 0.6266 |
| | FDD features | 0.5 s | 164.1 | 0.147 | 2.80 | 11 | 19.29 | **0.0228** |
| | FDD features | 1 s | 161.7 | 0.233 | 1.20 | 21 | 41.69 | **0.0020** |
| | FDD features | 5 s | 165.9 | 0.058 | 2.85 | 13 | 21.51 | **0.0285** |
| | FDD features | 10 s | 168.2 | 0.029 | 3.80 | 8 | 9.20 | 0.1628 |

The number of non-zero parameters in the model ($n_{params}$) including the intercept.
